# Study on the mucosal and serological immune response to the Novel Coronavirus (SARS-CoV-2) vaccines

**DOI:** 10.1101/2021.06.15.21256661

**Authors:** Renee WY Chan, Shaojun Liu, Jonathan Y Cheung, Joseph GS Tsun, Kate C Chan, Kathy YY Chan, Genevieve PG Fung, Albert M Li, Hugh Simon Lam

## Abstract

Vaccines that elicit mucosal immune responses against SARS-CoV-2 could potentially be of exceptional importance in providing first line defense at the site of viral entry. The serological antibody response induced by SARS-CoV-2 vaccines have already been well characterized. In order to understand the mucosal immune response profiles of SARS-CoV-2 vaccines, we examined both the mucosal and systemic responses of subjects vaccinated by two different vaccination platforms: mRNA (Comirnaty) and inactivated virus (CoronaVac). Serial nasal epithelial lining fluid (NELF) and peripheral blood samples were collected in ten subjects who had received CoronaVac and thirty-two subjects who had received Comirnaty. We quantified IgA and IgG specific to SARS-CoV-2 S1 protein by ELISA in NELF and plasma samples. The neutralization effect of these two sample types were evaluated by surrogate ACE-SARS-CoV-2 Spike protein ELISA. Only Comirnaty induced nasal SARS-CoV-2 S1 protein-specific (S1-specific) IgA and IgG responses, which were evident as early as on 14±2 days after the first dose. The NELF samples of 72% of subjects became IgA+IgG+, while in 62.5% of subjects the samples were neutralizing by 7±2 days after the second dose. In 45% of the subjects their NELF remained neutralizing 50 days after the booster of Comirnaty. In plasma, 91% and 100% Comirnaty subjects possessed S1-specific IgA+IgG+ on 14±2 days after the first dose and 7±2 days after booster, respectively. The plasma collected on 7±2 days after booster was 100% neutralizing. The induction of S1-specific antibody by CoronaVac was IgG dominant, and 70% of the subjects possessed S1-specific IgG by 7±2 days after booster and were all neutralizing. This study reveals that Comirnaty is able to induce S1-specific IgA and IgG response with neutralizing activity in the nasal mucosa in addition to a consistent systemic response. The clinical implications and the biological mechanism of an additional nasal immune response induced by vaccines such as Comirnaty warrant further investigation.

**One Sentence Summary:** mRNA vaccine (CoronaVac) elicits mucosal IgA and IgG in the nasal epithelial lining fluid together with ELISA-detected anti-wild-type spike neutralizing antibodies as early as day 14 post vaccination.

## INTRODUCTION

Severe acute respiratory syndrome coronavirus 2 (SARS-CoV-2) infection causes the coronavirus disease 2019 (COVID-19) pandemic that has resulted in significant morbidity and a global death toll of over 3 million (*1*).

Among the various COVID-19 vaccines currently authorized by the World Health Organization (WHO), CoronaVac by Sinovac Biotech and BNT162b2 (aka Comirnaty) from Pfizer-BioNTech have been approved for emergency use in Hong Kong. These two vaccines utilize different technological platforms, namely inactivated whole virus and messenger RNA (mRNA) encoding the full-length viral spike (S) protein modified by two proline mutations, respectively (*2*). The S-protein of SARS-CoV-2 is composed of the subunits S1 and S2, with S1 bearing the receptor-binding domain (RBD) that recognizes host angiotensin-converting enzyme 2 (ACE2) to initiate viral entry (*3, 4*) and S2 being responsible for membrane fusion (*3*). Both vaccines have good safety records with low prevalence of serious adverse events (*5-7*). CoronaVac has been shown to prevent symptomatic COVID-19 in 51% of vaccinated healthcare workers, and an efficacy of 100% in preventing severe COVID-19 (*8*). Comirnaty is reported to be 95% effective in preventing symptomatic COVID-19 with low incidence of serious adverse events (*5*).

Mechanistic properties of these novel vaccines in conferring immunity to combat COVID-19 are beginning to emerge. It has been shown that by 14 days after the booster dose, recipients of CoronaVac aged 18-50 years had seroconversion rates of 95.6% and 95.7% for S1-RBD IgG and neutralizing antibodies, respectively (*9*). In comparison, specific IgG against S1 and RBD of SARS-CoV-2 are detectable in serum at 21 days after the priming dose of Comirnaty, with 100% seroconversion rate (*10*). Due to the differences in the study design, it would be difficult to compare the seroconversion times for both SARS-CoV-2 S1 protein-specific (S1-specific) IgG and SARS-CoV-2 neutralization between the two vaccines. It is also important to note that due to differences in the stages of vaccine development, there is currently more research data available for the mRNA vaccines (e.g., Comirnaty and Moderna) than inactivated viral vaccines (e.g., CoronaVac).

Comirnaty and Moderna elicit neutralizing antibody (NAb) responses that target the RBD epitopes in the same manner as natural infections (*11*). Albeit at much lower titers than IgG levels, the mRNA vaccines also induce IgM and IgA responses against S-protein and RBD in plasma samples (*11*). In the sequence of seroconversion, IgM responses are first generated and then class-switch converted to IgA and IgG (*12*). As detectable IgM levels after vaccinations are often significantly lower and less sustained when compared to IgA and IgG levels, IgM is suspected to have lesser importance in virus neutralization *in vivo* (*11, 13*). On the other hand, serum RBD IgA from COVID-19 patients has been found to have more potent neutralization potential than paired IgG (*14*). SARS-CoV-2 IgA can be sustainably elevated in serum or plasma samples for over 2 months after Comirnaty vaccination (*13*). Thus, the importance of systemic SARS-CoV-2 IgA in vaccine-induced immunity against COVID-19 warrants further validation (*13, 15*).

Since SARS-CoV-2 infects the upper respiratory tract initially, local neutralizing antibodies could provide substantial protection against infection. In saliva of COVID-19 patients, SARS-CoV-2 IgA levels was found to be higher than IgG, with neutralization activity correlating with IgA titers, but not IgG (*14*). Natural infection induces mucosal antibodies directed against S-protein and RBD, and it has been suggested that higher antibody levels correlated with fewer systemic symptoms and reduced viral load (*16*). Thus, respiratory mucosal immunity could have unique and specific roles in offering protection against SARS-CoV-2 infection (*17*).

Secretory IgA (SIgA) is the most abundant immunoglobulin expressed on mucosal surface and serves as the first line of defense against infection (*14*). SIgA in the upper respiratory tract is from the IgA-secreting plasma cells (*18*). Whilst it is possible for systemic IgG-producing B cells to contribute the IgG in the respiratory tract, circulating monomeric IgA cannot be readily transported into secretions, suggesting that there are distinct systemic and mucosal responses to SARS-CoV-2 (*14*). Currently little is known about the mucosal immunity induced by SARS-CoV-2 vaccinations.

To date, one study has reported that mRNA vaccines induce detectable levels of salivary IgA and IgG responses to the S-protein and RBD of SARS-CoV-2, but their capability for viral neutralization are unknown (*19*). Determining whether or not current vaccines induce antibody response in the mucosa, and if so, the duration and sustainability of any such response in the context of concurrent data on systemic immunity from plasma samples of vaccinated individuals can provide invaluable information that can help optimize the use of these vaccines in a range of public health strategies and in different community settings.

The lack of sampling standardization and validation of the mucosal fluid measurements were the major challenges in the past. Our recent work using nasal strips to collect nasal epithelial lining fluid (NELF) in SARS-CoV-2 infected children and adults is non-invasive and facilitates the collection of undiluted NELF for SARS-CoV-2 nucleoprotein gene detection (*20*). The same method was adopted for the mucosal antibody analysis in the current study. With the possibility that subjects vaccinated with mRNA vaccine could induce salivary IgA against SARS-CoV-2 protein, we hypothesize that it might also induce SARS-CoV-2 S-protein specific antibody in other mucosal surfaces. Here, we compared the serological and mucosal immune responses after vaccination with CoronaVac and Comirnaty with a particular focus on S1-specific IgA levels detected and neutralization capacity in NELF and plasma.

## RESULTS

### Demographic of the subjects

Forty-two subjects were recruited in this study. The median age of all subjects was 41 years old (range 21-74), 40.3% were male. Longitudinal measurements of vaccine induced S1-specific IgG and IgA in serum and NELF at four time points (**Figure 1A**), 0 to 2 days before the first dose (Baseline), 14±2 days after the first dose (V+D14), 7±2 days after the booster dose (B+D7) and any time between 14 days after the booster dose and before 3 months after the first dose (**Figure 1A)** were conducted. The median age was significantly different in subjects from the two vaccine groups, CoronaVac (n=10, median age 59) and Comirnaty (n=32, median age 39.5) (*p=*0.0004). All subjects declared that they did not have known unprotected exposure to SARS-CoV-2 infected subjects. To ensure that the measurements of the change in SARS-CoV-2 specific immunoglobulin levels were not due to active SARS-CoV-2 infection during the study period, all the NELF samples were tested negative for the presence of SARS-CoV-2 nucleoprotein gene. As a result of the limited number of CoronaVac subjects and the detected difference in age, fifteen additional subjects who received CoronaVac were recruited for sampling at the 4th time point (**Figure 1B**). Among them, 43 subjects completed the questionnaire after vaccination to report any local or systemic events (**Supplementary Table 1**).

**Fig 1.**
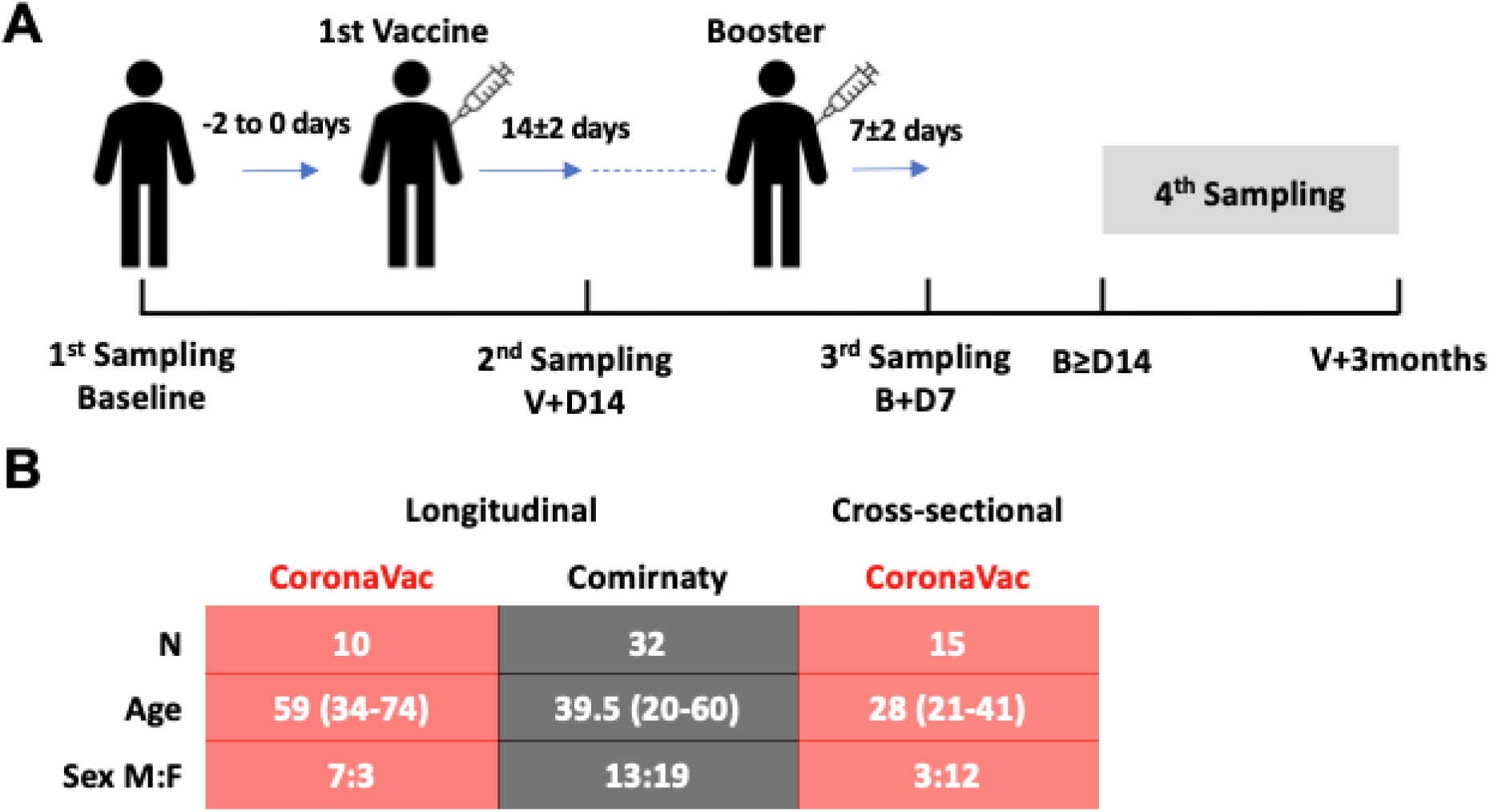
Study design and demographics. **(A)** There were three standard sampling timepoints and one extended sampling timepoint (4^th^ sampling) of biological sample collection: (i) 0 to 2 days before the first vaccination (Baseline), 14±2 days after the first vaccination (V+D14), (iii) 7±2 days after booster (B+D7) and (iv) any day between 14 days after booster and before 3 months after the first vaccination. **(B)** Subjects vaccinated with CoronaVac (n=10, pink table) and Comirnaty (n=32, grey table) were recruited and followed longitudinally, there was a significant difference in their age distributions (*p* = 0.0004, Mann Whitney test, two-tailed) and so fifteen extra subjects vaccinated with CoronaVac were recruited to enrich the data for the 4^th^ timepoint.

### Comirnaty induced detectable immunoglobulin in NELF

Among the ten subjects who have taken CoronaVac, none of them developed detectable NELF SARS-CoV-2 S1-specific IgA and IgG (**Figure 2A**) by day 7±2 days after booster, while most subjects who received Comirnaty developed S1-specific antibodies. The elevations in S1-specific IgA and IgG levels detected in NELF along the three time points were significant by two-way ANOVA followed by Tukey’s multiple comparisons test (**Figure 2D**). Moreover, S1-specific IgA appeared earlier than IgG in NELF. More subjects developed NELF S1-specific IgA 15/32 (46.9%) than IgG 3/32 (9.4%) **(Supplementary Figure 1B, blue dots)** by 14±2 days after the first vaccination. The proportion of subjects who had received Comirnaty were positive for NELF S1-specific IgA and IgG increased to 26/32 (81.3%) and 23/32 (71.9%), respectively, by 7±2 days after the booster dose **(Supplementary Figure 1C, blue dots)**.

**Fig 2.**
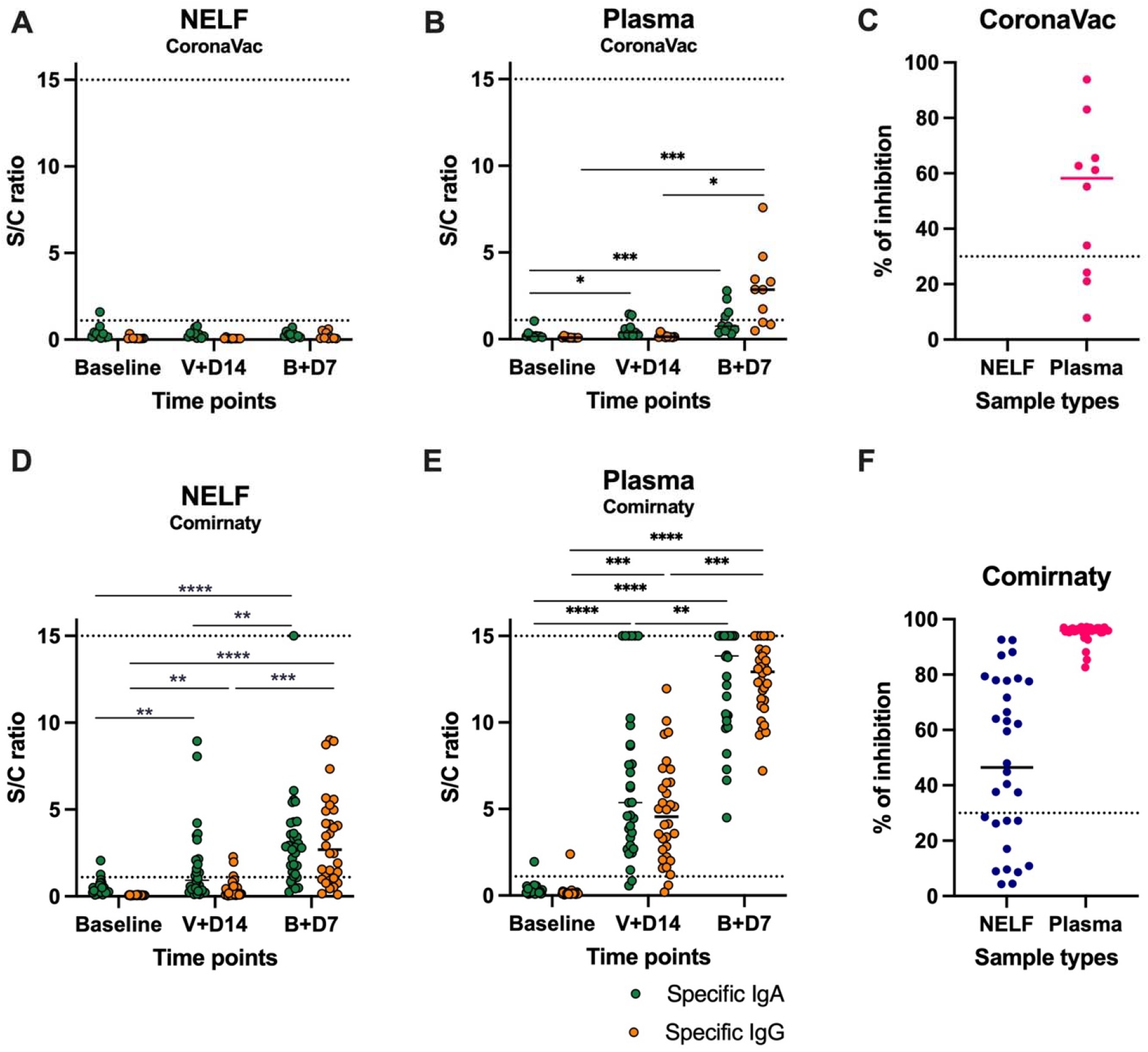
Expression of NELF and plasma SARS-CoV-2 S1-specific IgA and IgG and neutralizing antibody (NAb). The level of S1-specific IgA (green dots) and IgG (red dots) were plotted against the three standard timepoints of sample collection in NELF **(A and D)** and plasma **(B and E)** specimens of the recipients of CoronaVac **(A and B)** and Comirnaty **(D and E)**. Data points above the dotted line (Sample/Calibrator (S/C) ratio ≥ 1.1) are considered as positive, while the dotted lines at y=15 indicate the upper detection limit of the assay. Asterisks indicate statistical significance between timepoints of the same Ig class by two-way repeated-measures ANOVA followed by Tukey’s multiple comparison test. ^**^: *p*<0.001, ^***^: *p*<0.0002 and ^****^:*p*<0.0001. The percentage of signal inhibition observed with the surrogate SARS-Co-V 2 neutralization antibody detection kit by the NELF and plasma samples of CoronaVac **(C)** and Comirnaty **(F)** recipients collected on 7±2 days after booster were plotted. The 30% signal inhibition cutoff for SARS-CoV-2 NAb detection is interpreted as the sample containing neutralizing antibodies for SARS-CoV-2.

### SARS-CoV-2 IgA appeared earlier than IgG in plasma

In the CoronaVac group, plasma S1-specific IgA increased significantly by 14±2 days after the first vaccination dose and 7±2 days after the booster dose (**Figure 2B, green dots**), while the significant increase in S1-specific IgG was only detected between baseline and 7±2 days after booster (**Figure 2B, orange dots**). On 7±2 days after booster, 7/10 (70%) of the CoronaVac subjects had detectable plasma S1-specific IgG.

In the Comirnaty group (**Figure 2E**), 93.8% and 100% of subjects were positive for both plasma S1-specific IgA and IgG by 14±2 days after the first vaccination dose **(Supplementary Figure 1B,red dots)** and 7±2 days after the booster dose **(Supplementary Figure 1C, red dots)**, respectively. The outliers were contributed by three subjects who were IgA-IgG+ (Subject 40), IgA+IgG- (Subject 11) and IgA-IgG- (Subject 20). Nevertheless, all plasma samples were IgA+IgG+ by 7±2 days after booster.

There were no statistically significant correlations between age and the induced NELF and plasma S1-specific IgA and IgG levels in the longitudinal CoronaVac and Comirnaty recipients (**Supplementary Figure 2**). However, female subjects who received Comirnaty had higher plasma S1-specific IgG at the 14±2 days timepoint than male subjects (*p* = 0.007) (**Supplementary Figure 2D**).

### Neutralization potential of NELF and plasma

We further tested whether NELF and plasma samples were SARS-CoV-2 neutralizing by using the blocking enzyme-linked immunosorbent assay as a surrogate of the neutralization test. NELF and plasma samples collected at 7±2 days after booster were measured. In the CoronaVac group, as there were no SARS-CoV-2 S1-specific IgA and IgG detected in the NELF, we did not perform NAb measurement for those samples. Whereas 7/10 of the plasma from CoronaVac subjects contained SARS-CoV-2 NAb. In the Comirnaty group, 20/32 NELF samples inhibited the binding of SARS-CoV-2 spike protein to ACE-2 (**Figure 2F**), whereas all plasma samples provided over 80% inhibition to the binding of SARS-CoV-2 spike protein to ACE-2.

### The correlation between SARS-CoV-2 S1-specific Ig with neutralization antibody level

In the CoronaVac group, no significant correlations were found between the plasma IgA and IgG levels with the percentage of binding inhibition (**Figure 3A**). In the Comirnaty group, significant correlations were found between the S/C ratio of NELF IgA (r=0.509, *p*=0.031), NELF IgG (r=0.777, *p*<0.0001) (**Figure 3B**) and plasma IgA (r = 0.634, *p*=0.008) levels and the percentage of binding inhibition (**Figure 3C**).

**Fig 3.**
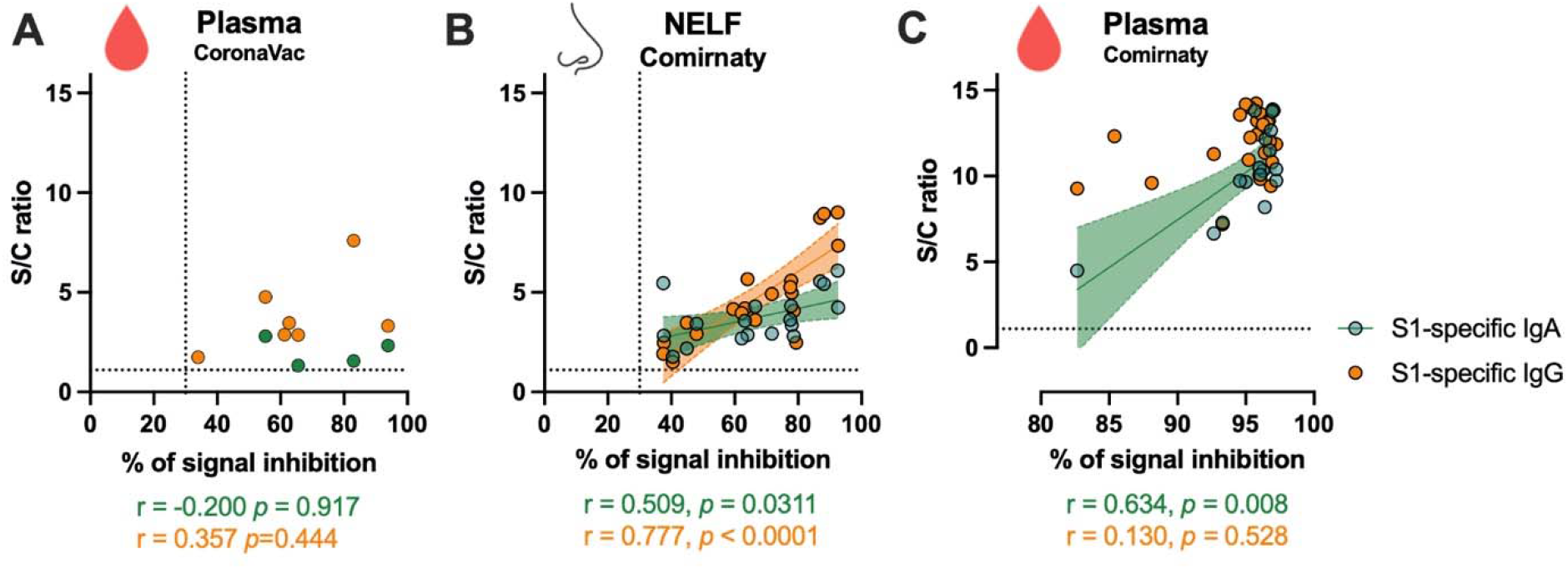
Correlation of SARS-CoV-2 S1-specific Igs to the percentage of signal inhibition in the surrogate ACE-2 based neutralization readout. The correlation coefficients of the S/C ratio of the **(A)** plasma of CoronaVac subjects, **(B)** NELF and **(C)** plasma of Comirnaty subjects at 7±2 days after booster are superimposed on the panel with trend lines estimated with the use of simple linear regression. Plots show the S/C ratio of the SARS-CoV-2 S1-specific IgA (green dots) and IgG (orange dots) between ≥1.1 to <15 were plotted against the percentage of inhibition of the SARS-CoV-2 Spike-ACE-2 binding signal, in which an inhibition ≥ 30% is regarded as the threshold of a positive sample, indicated by the vertical dotted line. Data with negative result in either one test were excluded from the two-tailed Spearman correlation analysis. Green and orange dotted lines represent significant linear regression fits with 95% confidence intervals (shaded region with the corresponding colors).

### Neutralizing antibody in NELF is transient

Whether this is a transient expression SARS-CoV-2 NAb in NELF is uncertain, as there were five individuals (subjects 9, 33, 34, 37, 41) showing a downward trend of S/C ratio of IgA from 14±2 days after the first vaccination to 7±2 days after booster (**Supplementary Figure 2A**). The longevity of the NAb in NELF was further assessed in 11/32 Comirnaty subjects who had reached the 4^th^ sampling time point, i.e. any time after 14 days post-booster and before 3 months post-vaccine (**Figure 4A**). Only three NELFs still contained NAb, while two of them became NAb negative. Nevertheless, a late NELF NAb development was observed in two individuals, who did not possess NELF NAb earlier on 7±2 days after booster, though four subjects’ NELF remained negative for NAb. Within these eleven subjects, a significant decrease in the S/C ratio of the S1-specific IgA was also observed from 7±2 days after booster to the 4th sampling time point (*p*=0.018) (**Supplementary Figure 3**). Lastly, at the 4th sampling time point, the 25 NELFs obtained from the CoronaVac group also underwent the NAb assay, however, none of them were positive, though one subject from the cross-sectional group had a positive S1-specific IgA (S/C ratio=1.27) readout.

**Fig 4.**
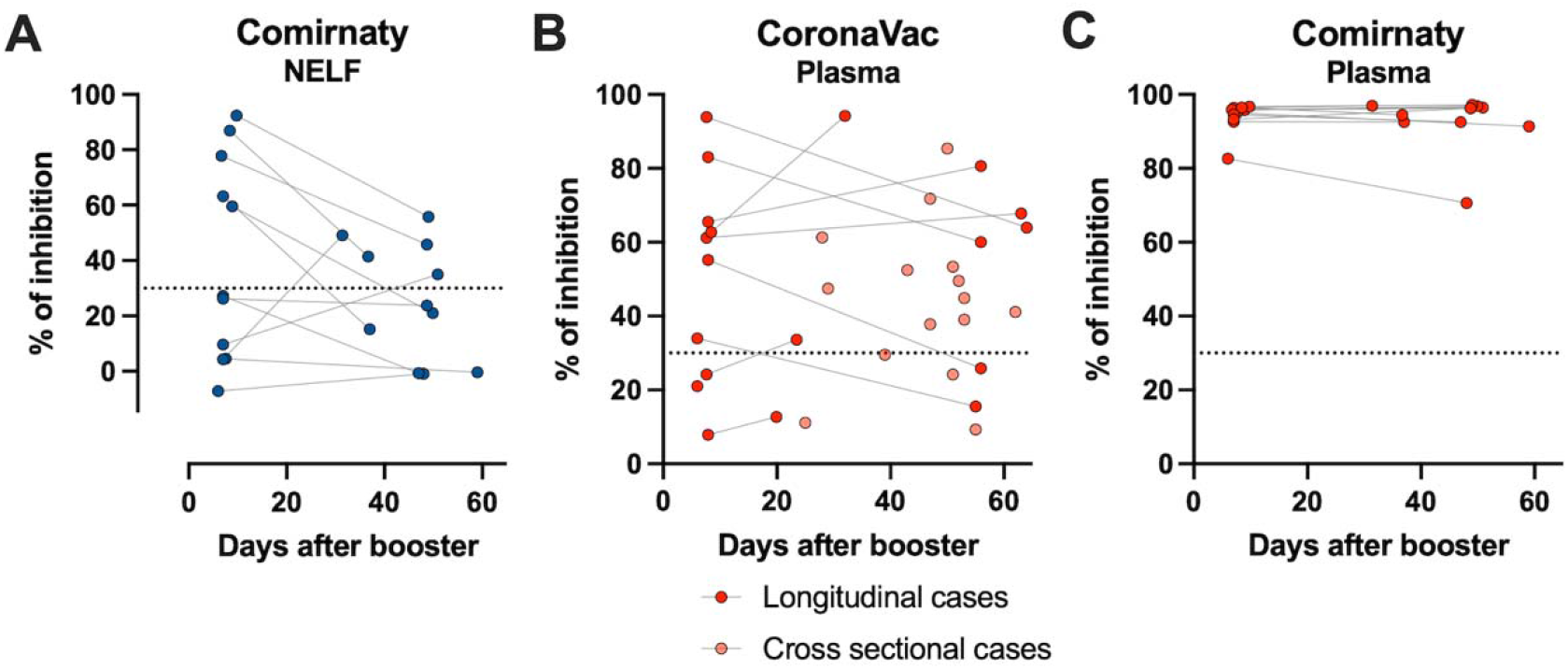
Longevity of the NAb in NELF and plasma samples. **(A)** The paired-percentage of signal inhibition by the NAb in the NELF from 7±2 days after booster to the 4th sampling time point of eleven Comirnaty subjects. Data from the samples of the same individual is joined by a dotted line. **(B)** The paired-percentage of signal inhibition by NAb in the plasma from 7±2 days after booster to the 4th sampling time point of CoronaVac subjects in the longitudinal group (n=9, red dots) is joined by a dotted line, and each pink dot represents the percentage of signal inhibition of the plasma samples from CoronaVac recipients in the cross-sectional group (n=15). **(C)** The paired-percentage of signal inhibition by NAb in the plasma of eleven Comirnaty subjects from 7±2 days after booster to the 4th sampling time point is shown.

### 75% NAb detection in CoronaVac subjects’ plasma

After 7±2 days of the booster 7/10 subjects who had received CoronaVac had NAb (**Figure 4B**), and five of them remained NAb positive by their 4th sampling time point. With the extended time after booster, one gained NAb by 24 days after the booster (subject 13) though another subject did not develop plasma NAb even by 20 days post booster (Subject 14). The differences in observations in our longitudinal cohort of Sinovac recipients (n=9) from Comirnaty recipients were unlikely an age effect, as we found that consistent with the longitudinal subjects, 12/15 plasma samples from the younger subjects in the cross-sectional group of CoronaVac recipients had NAb (**Figure 4B, pink dots**). Overall, 75% (18/24) of the recruited subjects had NAb in their plasma at the 4th sampling time point. Moreover, there were no correlations between NAb levels either with time post booster (p=0.38) or with age (p=0.78). In the Comirnaty group, all subjects had NAb since 7±2 days after booster and it lasted at least for 50 days after the booster dose (**Figure 4C**).

## DISCUSSION

Our study reveals that both Comirnaty and CoronaVac induces plasma SARS-CoV-2 S1-specific IgA and IgG, and NAb. However, Comirnaty, but not CoronaVac, induced S1-specific IgA and IgG in the nasal mucosa by 7±2 days after booster. Of the Comirnaty recipients 72% produced an NELF antibody response, while 62.5% exhibited neutralizing activities in their NELF samples. The NAb in NELF correlated with the S/C ratio of the S1-specific IgA and IgG detected. The induction of nasal S1-specific Igs and NAb is unique to subjects receiving Comirnaty and it was not found in the NELF of the CoronaVac recipients. The longevity of the NAb in NELF was assessed in 12/32 Comirnaty subjects who had reached the extended time point, i.e. any day between 14 days after booster and before 3 months after the first vaccination. Only three NELFs remained neutralizing, while two of them became NAb negative. Nevertheless, a late rise in NELF NAb was observed in two individuals who did not possess NAb in their NELF in the earlier time points. Lastly, four of the Comirnaty recipients remained NELF NAb negative through all timepoints. From our data, the plasma NAb may last at least 50 days after the booster, though further samples are required to validate this claim.

It has been commonly believed that intramuscular vaccines do not induce mucosal immunity effectively (*21*). The mucosal immune response of the upper respiratory tract is partly compartmentalized and usually initiated in nasopharynx-associated lymphoid tissue (NALT) in all age groups and bronchus-associated lymphoid tissue (BALT) in children and adolescents or in adults upon disease induction (*18*). These upper respiratory tract associated lymphoid tissues generate IgA-producing mucosal B cells that express the homing receptor, e.g. α4ß1, CCR10, CD62L and LFA-1 (*22, 23*). These homing receptors allow the B-cells to traffic efficiently to the mucosal effector site, the respiratory tract in this case, where their ligands VCAM-1 and CCL28 are strongly expressed. The IgA-producing mucosal B cells differentiate into polymeric IgA-secretory plasma cells and contribute to the production of the polymeric IgA (in dimers or tetramers) in the lamina propria as opposed to serological IgA (predominantly monomers), which is produced within bone marrow, spleen and lymph nodes (*17*). Therefore, SIgA present in secretions are typically produced within mucosal tissues. This raises important questions about the route that mRNA-lipid nanoparticles would take from the intramuscular injection site to the NALT (and BALT) and the biological mechanisms that underlie this process.

In an animal study using similar lipid nanoparticles carrying mRNA encoding haemagglutinin proteins of influenza to investigate the biodistribution of the influenza mRNA in mouse plasma and tissue after intramuscular administration. It is assumed that the concentration of mRNA lipid nanoparticles decreases along the disseminating route from the injection site which includes through the systemic circulation and via the lymphatic system, spleen and liver. It was found that the expression of mRNA can be detected in distal tissues, including lung, though the concentration was 1,000 fold lower (*24*). The same study also showed the presence of mRNA in the intestine, though they did not test for the presence of such in the airway mucosae. In terms of the Comirnaty vaccine, the 30-microgram vaccine dose contains approximately 13,000 billion repetitions of the optimized SARS-CoV-2 Spike protein sequence as documented (*2*). We postulate that the number of the mRNA lipid-nanoparticles that reach the nasal mucosa after Comirnaty injection might be sufficient for NALT stimulation. However, the mechanisms underlying this process and factors that affect the consistency of this effect requires further investigation.

The clinical implication of the induction of nasal SARS-CoV-2 NAb is increased likelihood of immediate protection at the target site of viral infection. The role that this mucosal immune response may play in reducing the risk of virus transmission should also be considered. Sufficiently high concentrations of NAb in the NELF can block the attachment of virus to the host cell receptor. The SIgA is known to trap virus efficiently and facilitates the removal of such by the mucociliary beating (*25*), to neutralize virus particles with epithelial cell and lamina propria (*17*), and has anti-inflammatory properties (*26*).

We also confirmed the presence of vaccination induced plasma S1-specific IgA and IgG in both CoronaVac and Comirnaty recipients. For CoronaVac, 70% of the subjects were found to have NAb by 7±2 days after booster. Whereas all subjects who received two doses of Comirnaty had NAb in their plasma samples 7±2 days after booster. Of note, at the 4th sampling time point, 73% of the cross-sectional CoronaVac group had plasma NAb (**Figure 4A**, pink dots), which is similar to the older subjects in the longitudinal CoronaVac group, inferring that age is not a contributing factor to the NAb level in the CoronaVac group. Therefore, the lower percentage of plasma NAb detected in CoronaVac than the Comirnaty group cannot simply be explained by the difference in age.

The delayed NAb response found in CoronaVac subjects compared with those in Comirnaty group was not surprising. It has been reported that seroconversion rates were 47.8% and 95.6% for S1-RBD-specific IgG for CoronaVac at 14 days and 28 days after booster, respectively (*9*). Thus, our study design with this interim report at 7 days ± 2 post booster may not have demonstrated the full immune responses elicited by CoronaVac. Nevertheless, there were seven CoronaVac subjects who did not develop NAb in their plasma samples even by the 4th sampling time point. Three of them were from the longitudinal group in which two of them had their NAb weaned on 55 and 56 days after booster (Subjects 8 and 5, respectively) while one of them never exhibited plasma NAb even after 19 days of booster (Subject 14). For the four NAb negative plasma results contributed by the cross-sectional group, without the baseline measurements, we cannot conclude whether this was due to a short duration of their NAb responses or if they never developed a NAb response. The apparent non-responders were subjects L7, L25, L37 and L38 at their 55, 51, 39 and 25 days after booster, respectively. This might infer the needs of an additional dose to ensure a sufficient protection to the CoronaVac recipients (*27*). Moreover, apart from the humoral response towards spike protein, the development of antibodies against other viral proteins which have a lower mutation rate, such as nucleoprotein, might be an edge of CoronaVac over Comirnaty (*28*). Furthermore, a comprehensive measurement to include the cellular response induction would provide a balanced information about the overall protection against SARS-CoV-2 exerted by vaccination (*29*).

Consistent with our findings, Danese *et al*. demonstrated that all SARS-CoV-2 antibodies (IgM, IgG, IgA) begin to rise from 7 to 11 days post primer dose of Comirnaty and they also showed that the booster dose of Comirnaty further increases the levels of IgG against S1/S2 and RBD (*13*). Both plasma IgG and IgA levels have been found to remain elevated for up to 65 days post first vaccine dose (*13*). Whilst Wang *et al*. reported that after receiving two doses of mRNA vaccines (Comirnaty or Moderna), high levels of IgM and IgG against S and RBD of SARS-CoV-2 is detectable for up to 8 weeks after booster (*11*). Furthermore, our study demonstrated the correlation between plasma IgA levels with the percentage of virus-receptor binding inhibition as reported in a previous study (*14*). Together, our findings confirm the reliability of Comirnaty in generating robust humoral immune responses in vaccinated subjects.

The presence of nasal mucosal immunoglobulins after vaccination against COVID-19 has not been previously reported. Whilst we currently have some insights into the durability of serological IgA and IgG response after Comirnaty vaccination. There is currently no information on the longitudinal expression of the immunoglobulins in NELF samples representing mucosal immunity in COVID-19 patients nor recipients of SARS-CoV-2 vaccine. We are now continuously following these subjects and collecting paired NELF and blood samples at 3, and 6 months after first dose of vaccination to better understand the longevity of mucosal immunity elicited by intramuscular vaccination. Such findings could have implications on public health strategies and screening for immunity to enable resumption of normalcy on a global scale.

The current study has the following limitations. First, the smaller sample size and the higher median age of the recipients of CoronaVac in the longitudinal group can be argued to have contributed towards the absence in NELF response and a slower and milder plasma response when compared to Comirnaty. We attempted to recruit the cross-sectional subjects to enrich our data for this important early report, while more subjects would be recruited for a better comparison. Second, we were using a SARS-CoV-2 surrogate virus neutralization test instead of a neutralization assay with live cells and viruses in Biosafety Level 3 settings. Therefore, the NAb measured in this study is a surrogate measure that is solely based on the inhibition of the binding between the SARS-CoV-2 antibody-mediated blockage of ACE2-spike (RBD) protein-protein interaction (*30*). The protective effects of the intracellular action of NELF IgA in the Comirnaty recipients or the plasma Ig specific to other SARS-CoV-2 proteins that theoretically should be manufactured in CoronaVac recipients were not considered. Although the surrogate assay used has been validated with the plaque reduction neutralization test (PRNT) utilizing the SARS-CoV-2 virus (*31*), the current test could underestimate the actual neutralization capacity. Third, we observed tremendous individual variations, for example, some recipients of Comirnaty were found to be IgA-, IgG- or IgA-IgG- for S1 protein. Although Comirnaty induced mucosal immunoglobulins for 72% of vaccinated subjects, there were individuals who, despite having plasma S1-specfic IgA and IgG, did not express NELF S1-specific IgA (n=2) or IgG (n=5) or both (n=4). These variations require a larger sample size to further clarify. Nevertheless, our current study clearly shows qualitative and significant differences in mucosal response between different vaccine technologies. Lastly, the best available assay for the measurement of SARS-CoV-2 S1-specific IgA and IgG used in this study was optimized for plasma and serum rather than mucosal lining fluid. We were therefore unable to differentiate between the monomeric and dimeric forms of IgA or identify any secretory component in our subjects’ NELF samples.

## CONCLUSION

Despite being a vaccine administered via the intramuscular route, Comirnaty, and likely other mRNA vaccines, uniquely induces SARS-CoV-2 S1 specific IgA and IgG in the nasal mucosal of vaccine recipients as early as 14 days after the first dose. The NELF neutralizing effect infers protection from SARS-CoV-2 infection at the level of the upper respiratory epithelium, when the level of NAb is sufficiently high. This extra arm of protection at the mucosa, on top of the well-characterized serological antibody development, might further reduce the chance of SARS-CoV-2 infection, in addition to its effectiveness in protecting the recipient from hospitalization and severe disease. Though the response may be transient, it is possible that a more rapid elevation in antibodies may occur within the mucosa when the subject is exposed to live viruses, thus conferring protection from infection even before the virus breaches the mucosa. CoronaVac vaccine induces an IgG dominant response in the recipients’ plasma with neutralizing effect, but did not produce any mucosal antibody response. The duration required for the plasma Igs and NAb development observed in our study, is comparable to what has been previously reported, however, the additional information relating to mucosal response and the direct comparison between two vaccine technologies provides important insights into how to best utilize these different vaccines from a public health point of view.

## IMPLICATIONS

This study provides paired data of the mucosal findings and systemic immunological endpoints of adults before and after receiving the two different SARS-CoV-2 vaccines. The insights gained from the different immune profiles between inactivated viral vaccines and mRNA vaccines would be helpful to help optimize public health strategies. While we found that both Comirnaty and CoronaVac induced systemic humoral responses, Comirnaty likely provides enhanced mucosal level immune protection and which we postulate could contribute to reduction in asymptomatic transmission risk. This suggests that Comirnaty may be more suitable for individuals who are often in close contact with vulnerable and/or unvaccinated individuals (e.g., old age home workers, pediatricians, school teachers).

CoronaVac, on the other hand, induced satisfactory systemic humoral response with neutralizing capacity induced by CoronaVac in most individuals, and there is less concern about the potential for unintended inflammatory or immune reactions in organs/tissues distal to the vaccination site means that CoronaVac may be more suitable for large groups of vulnerable populations who require protection from SARS-CoV-2. The easier logistics involves in the storage and distribution of CoronaVac might help to provide a high vaccination coverage within such vulnerable populations.

The unexpected mucosal response in mRNA vaccine recipients raises the concern about which other organs/tissues may be similarly affected and whether inflammatory/immunological responses in some tissues may cause unintended side effects with adverse outcomes. Therefore, this piece of information highlights the necessity in speeding up further studies to determine the distribution of mRNA lipid nanoparticles in humans. Moreover, the mucosal humoral response against SARS-CoV-2 S protein is inconsistent between individuals. The publication of this manuscript will consolidate the collaborative effort among different research groups to investigate the biological determinant to increase the consistency of mucosal response.

Finally, we see a niche for the further development of the antibody detection strategy by using mucosal lining fluid sample. The current collection method is painless, self-administered and can be carried out repeatedly in all age groups. The mucosal antibody test would provide the immune status after vaccination (or by natural infection), with a direct reflection of the protection level at the site of virus entry.

## MATERIALS AND METHODS

### Subject recruitment

We released the information of this study through the department website, department social media, The Chinese University of Hong Kong (CUHK) mass mail system, CUHK-online form and word of mouth to reach potential subjects and arrange the sampling logistics. Subject who had arranged their own COVID-19 vaccinations with known schedules of vaccine doses were recruited. All subjects were requested to complete a one-page questionnaire to capture their demographics, past medical history, drug use and the reporting of any adverse effects after vaccination or respiratory tract infections within the study period. Consent was obtained from the participants and the study was approved by the Joint Chinese University of Hong Kong – New Territories East Cluster Clinical Research Ethics Committee (CREC: 2021.214).

### Clinical sample collection regime

Nasal lining epithelial lining fluid (NELF) from both nares and 3mL peripheral blood were collected from the subjects at four time points. A pre-vaccination sample pair was collected during the 48-hour period before the day of vaccination. Three post-vaccination sample pairs were collected at 14 ± 2 days after first dose and 7 ± 2 days post-booster as in **Figure 1A**. An extended sampling timepoint (4^th^ sampling) of biological sample collection was performed any day between 14±2 days after the booster and before 3 months after the first vaccination dose, to assess the intermediate longevity of SARS-CoV-2 specific Ig and NAb responses.

### NELF collection by nasal strips

Strips were cut from sheets of Leukosorb medium (Pall Corporation, BSP0669) using a laser cutter (CMA960, Department of Biomedical Engineering, CUHK) to the dimensions of 4 mm wide and 40mm long with a marking at 12mm as previously described (*20, 32*). 100uL of sterile saline were instilled in each nostril of the subject. Strips were inserted into the anterior part of the inferior nasal turbinate of each nostril until the indicator mark was at or close to the base of each naris. After insertion, the nose was pinched for 1 minute to allow thorough absorption of NELF by the strip. Strips were removed and eluted within 24h after collection. To elute NELF, strips were soaked in 300uL PBS on ice for 5 min with a quick vortex. The solution and the strip were transferred to a Costar Spin-X (CLS9301) and centrifuged twice at 13,000 rpm for 2 min at 4°C to elute the NELF from the strip into the 1.5-ml tube. The NELF was aliquoted into small volume vials for downstream analysis of SARS-CoV-2 specific Ig panels and neutralization test and were stored at −80°C until analysis.

### Plasma preparation

Blood was collected aseptically by venepuncture and transferred into EDTA blood tube. Plasma samples were separated by centrifugation at 4°C, 2000g for 20 minutes, aliquoted into small volume specimens and stored at –20° C until analysis.

### Measurement of specific IgA and IgG against SARS-CoV-2 Spike protein

Semi-quantitative measurements of SARS-CoV-2 Spike protein (S1 domain) specific Ig ELISA Kits (Euroimmun, EI 2606-9601 A and EI 2606-9601 G) were used. 1:10 diluted NELF and 1:100 diluted plasma were added to the assay well and processed as per manufacturer’s instructions. After subsequent wash and incubation steps with conjugates or substrates, the plates were analyzed according to the manufacturer’s instructions on the Synergy HTX Multi-Mode Reader. Semi-quantitative readout as a ratio between the sample and the calibrator optical density (OD) values was used. The performance was checked by keeping the optical density of the calibrator within the reference value, and the ratio between the positive and negative controls between 1.6-4.2 and 0-0.7, respectively.

### Measurement of SARS-CoV-2 neutralization antibody

A blocking enzyme-linked immunosorbent assay (GenScript, L00847) was employed. Briefly, NELF, plasma samples and controls were 1:9 diluted and mixed with HRP-RBD solution and incubated at 37°C for 30 minutes. The mixture was then added to the human ACE-precoated plate and incubated at 37°C for 15 minutes and processed as per manufacturer’s instructions. The performance was checked by ensuring that the OD450 must fall below 0.3 for the positive control and above 1.0 for the negative control. A 30% signal inhibition was set as the cutoff for SARS-CoV-2 NAb detection.

### Viral RNA extraction and quantification

To eliminate the possibility of active SARS-CoV-2 infection during the study period, 70uL of NELF collected at each timepoint were extracted using PHASIFY VIRAL RNA Extraction Kit™ following manufacturer’s instruction. RNA was reconstituted in 20uL of RNase-free water. 4uL of the RNA sample was used in each reaction, and the SARS-CoV-2 RNA was quantified by one-step Master Mix (TaqMan Fast Virus, ThermoFisher) with primers and probe targeting the N gene of SARS-CoV-2 as described (*33*). Duplicate reaction was conducted on QuantStudio 12K Flex Real □Time PCR System (Applied Biosystems, Foster City, CA, USA) at the following cycling conditions: reverse transcription at 50°C for 5□min, inactivation of reverse transcriptase at 95°C for 20□s, 40 cycles of PCR amplification (Denaturing at 95°C for 5□s; Annealing/ Extending at 60°C for 30□s). No template control and positive control using cell lysate from SARS-CoV-2 infected human respiratory cells were included in each run.

### Statistical analysis

The demographic variables of subjects were compared between the two vaccinated groups using the Mann Whitney test and Fisher’s exact test, as appropriate. For the immunoglobulin profiles, differences between different gender and time points were evaluated using Mann Whitney test and Friedman test followed by Dunn’s multiple comparison test, respectively. The correlation of S/C ratio of the specific immunoglobulins with the percentage of signal inhibition in the surrogate neutralization test was examined by Spearman’s correlation test. Differences were considered to be statistically significant if *p* < 0.05. The All statistical tests were performed using Graphpad version 9.1.2 for macOS. Differences were considered statistically significant at *p* < 0.05.

## Data Availability

The raw data will be available upon request.

## Acknowledgments

We would like to acknowledge Prof Aaron HP Ho, Prof Megan YP Ho and Miss Yuan-yuan Wei (Department of Biomedical Engineering, Faculty of Engineering, The Chinese University of Hong Kong) who tailor-cut the nasal strips for this study and Ms Fiona Cheng (Department of Paediatrics, The Chinese University of Hong Kong) for her assistance in preparing all the nasal strip vials. We would like to offer our special thanks to Ms Carrie Lee and Ms Cecily Leung in collecting the biospecimens from our research subjects and all the subjects who agreed to participate in this study. We thank Dr Agnes Leung, Prof Tony Nelson and Prof Ellis KL Hon (Department of Paediatrics, The Chinese University of Hong Kong) for their continuous encouragement to the research team.

## Author contributions

Conceptualization: KYYC, HSL, RWYC

Methodology: SL, JGST, KYYC, HSL, RWYC

Investigation: SL, JGST, KCCC, KYYC, GPGF, HSL, RWYC

Visualization: SL, JYC, JGST, KYYC, RWYC

Funding acquisition: HSL, AML, RWYC

Project administration: SL, JGST, KYYC, RWYC

Supervision: KYYC, HSL, AML, RWYC

Writing – original draft: SL, JYC, RWYC

Writing – review & editing: KCCC, KYYC, HSL, AML, RWYC

