## Supplementary Appendix for "Study on the mucosal and serological immune response to the Novel Coronavirus (SARS-CoV-2) vaccines"

Renee Wan Yi Chan, PhD, MPhil, BSc

Assistant Professor, Department of Paediatrics, Faculty of Medicine, The Chinese University of Hong Kong

Corresponding Address: Lab D, 8/F Tower A, The Hong Kong Children's Hospital, Kowloon Bay, Kowloon, HONG KONG

Office Phone: 852+3513-3175

Office Fax: 852+2636-0020

Hugh Simon Lam,

BChir (Cantab), MB (Cantab), MD (CUHK), MRCPCH (UK), FHKCPaed, FHKAM (Paediatrics), FRCPCH (UK)

Professor, Department of Paediatrics, Faculty of Medicine,

The Chinese University of Hong Kong

Corresponding Address: 6/F Lui Chee Woo Clinical Sciences Building

Office Phone: 852+3505-2851 Office Fax: 852+2636-0020

### Supplementary Results

#### Local and systemic events after vaccination

In the CoronaVac cross-sectional group (n=15), all fifteen subjects, who have already received two doses of the vaccine, filled in a questionnaire to report their past medical history, as well as local and systemic events after vaccination (**Supplementary Table 1A**). No subjects reported history of allergic disease. One subject has hyperlipidemia and was not on any drug treatment. 5/15 (33%) subjects experienced injection site pain. No subjects reported systemic events.

In the Comirnaty longitudinal group (n=32), twenty-eight completed questionnaires were received from twenty-three subjects, of whom five completed the survey after two doses of Comirnaty, seven after the first dose only, and eleven after the booster only. Thus, twelve subjects completed their questionnaires after the first dose and 16 subjects filled in the same questionnaire after the booster. From the sixteen subjects who have received both doses of Comirnaty, nine reported their past medical history of allergic diseases, including allergic rhinitis (31%), asthma (6%), eczema (6%), food allergy (19%) and drug allergy (6%). Three subjects had either hypertension (n=1) or hyperlipidaemia (n=2). 11/16 (69%) subjects experienced injection site pain and one reported injection site swelling. No subjects reported systemic events. A significantly higher percentage of Comirnaty subject experienced muscle pain than those reciving CoronaVac ($p < 0.01$) (**Supplementary Table 1A**).

#### Comparing effects after first and second dose of Comirnaty

10/12 and 11/16 subjects reported local events after the first dose and the booster, respectively. The local events included injection site pain (83% vs 69%), redness (17% vs 0%) and swelling (25% vs 6%). 7/12 and 13/16 subjects reported systemic events after the first and second dose respectively, including muscle pain (42% vs 44%), fatigue (33% vs 44%), fever (8% vs 25%), headache (8% vs25%), drowsy (0% vs 13%), loss of appetite (0% vs 13%), limb pain (8% vs 6%), limb numbness (8% vs 6%) and sore throat (0% vs 6%) (**Supplementary Table 1B**).

| | CoronaVac Cross-sectional | | Comirnaty Longitudinal | | ***P* value** |
|---|---|---|---|---|---|
| **Subjects** | N=15 | | N=16 | | |
| **Age (mean and range)** | 29.3 (21-41) | | 40.3 (21-57) | | **<0.01** |
| **Gender (male:female)** | 3:12 | | 9:7 | | 0.07 |
| **BMI** | 19.72 (17.4-22.8) | | 23 (18.59-33.10) | | **<0.01** |
| **Allergic disease** | | | | | |
| Allergic rhinitis | 0 | 0% | 5 | 31% | **0.04** |
| Asthma | 0 | 0% | 1 | 6% | >0.99 |
| Eczema | 0 | 0% | 1 | 6% | >0.99 |
| Food allergy | 0 | 0% | 3 | 19% | 0.22 |
| Drug allergy | 0 | 0% | 1 | 6% | >0.99 |
| **Other disease** | | | | | |
| hyperlipidemia | 1 | 7% | 1 | 6% | >0.99 |
| hypertension | 0 | 0% | 2 | 13% | 0.48 |
| **Local event** | | | | | |
| injection site pain | 5 | 33% | 11 | 69% | 0.07 |
| injection site redness | 0 | 0% | 0 | 0% | - |
| injection site swelling | 0 | 0% | 1 | 6% | >0.99 |
| **Systemic side effect** | | | | | |
| muscle pain | 0 | 0% | 7 | 44% | **<0.01** |
| fatigue | 2 | 13% | 7 | 44% | 0.11 |
| fever | 0 | 0% | 4 | 25% | 0.10 |
| headache | 0 | 0% | 2 | 13% | 0.48 |
| drowsy | 0 | 0% | 2 | 13% | 0.48 |
| loss of appetite | 0 | 0% | 2 | 13% | 0.48 |
| chill | 0 | 0% | 1 | 6% | >0.99 |
| limb pain | 0 | 0% | 1 | 6% | >0.99 |
| **Contact with Covid-19 patients** | 0 | 0% | 0 | 0% | - |
| **Positive for SARS-CoV-2 test ever** | 0 | 0% | 0 | 0% | - |

**Table S1A. Subject demographics, medical history and the local and systemic side effects reported in the CoronaVac and Comirnaty group after booster.** Fisher's exact test (two-tailed) was performed to determine the difference in the medical history, local and systemic side effect between the two vaccine groups.

| | After first dose of Comirnaty (n=12) | | After booster of Comirnaty (n=16) | | *P* value |
|---|---|---|---|---|---|
| **Local side effect** | | | | | |
| injection site pain | 10 | 83% | 11 | 69% | 0.66 |
| injection site redness | 2 | 17% | 0 | 0% | 0.17 |
| injection site swelling | 3 | 25% | 1 | 6% | 0.29 |
| **Systemic side effect** | | | | | |
| muscle pain | 5 | 42% | 7 | 44% | >0.99 |
| fatigue | 4 | 33% | 7 | 44% | 0.70 |
| fever | 1 | 8% | 4 | 25% | 0.15 |
| headache | 1 | 8% | 2 | 13% | >0.99 |
| sleepy | 0 | 0% | 2 | 13% | 0.49 |
| loss of appetite | 0 | 0% | 2 | 13% | 0.49 |
| chill | 0 | 0% | 1 | 6% | >0.99 |
| limb pain | 1 | 8% | 1 | 6% | >0.99 |
| limb numbness | 1 | 8% | 0 | 0% | >0.99 |
| sore throat | 0 | 0% | 1 | 6% | >0.99 |

**Table S1B. Local and systemic side effects reported in the Comirnaty group after the first dose and booster.**

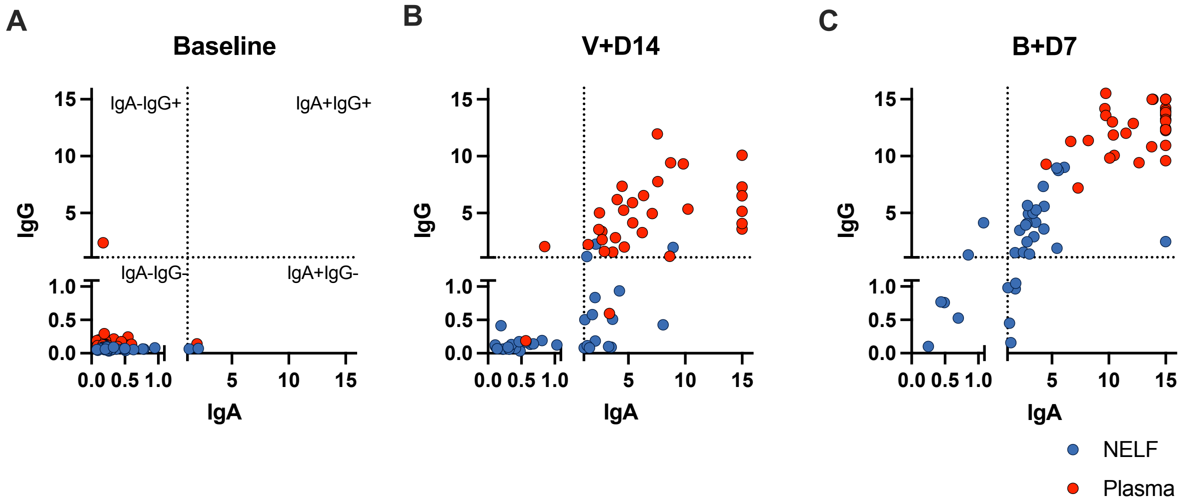


**Fig S1. Dynamics of IgA and IgG in NELF and plasma of subjects received Comirnaty.** The level of the SARS-CoV-2 S1-specific IgG in NELF (blue dots) and plasma (red dots) were plotted against IgA at **(A)** baseline, **(B)** on 14±2 days after the first dose of Comirnaty and **(C)** 7±2 days after booster. The dotted lines represent the positive thresholds of S1-specific IgA and IgG of the assays, and the dots fall within the four areas represent their nature in having IgA-IgG+, IgA+IgG+, IgA-IG- and IgA+IgG- as illustrated in **(A)**.

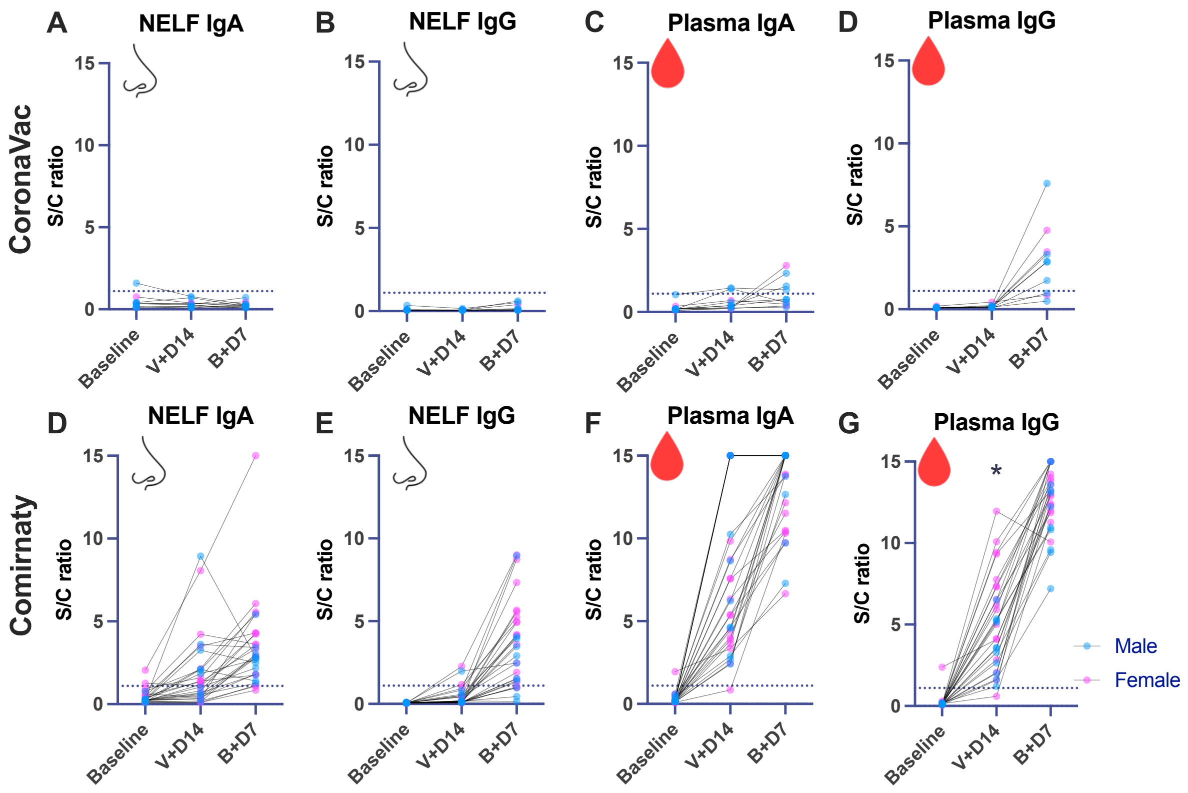


**Fig S2. Female recipients of Comirnaty had a higher SARS-CoV-2 S protein specific IgG in plasma on 14±2 days after the first vaccination.** The level of the SARS-CoV-2 S1-specific **(A,D)** IgA in NELF, **(B,E)** IgG in NELF, **(C,F)** IgA in plasma and **(D,G)** IgG in plasma were plotted against the three timepoints of sample collection according to male (n=7; n=11, blue dots) and female (n=3; n=17, pink dots) of the longitudinal **(A-D)** CoronaVac and **(D-G)** Comirnaty group, respectively. Outliers who were unable to produce detectable SARS-CoV-2 S1-specific IgA in NELF at all time points in Comirnaty group were removed (Subjects 16, 20, 22 ,48). The lines connected the Ig levels detected in the same subjects at different time points. Data points above the dotted line (Sample/Calibrator ratio ≥ 1.1) are considered as positive. Asterisks indicate statistical significance between genders at a specific time point with ($p$ = 0.0417, Mann-Whitney test).

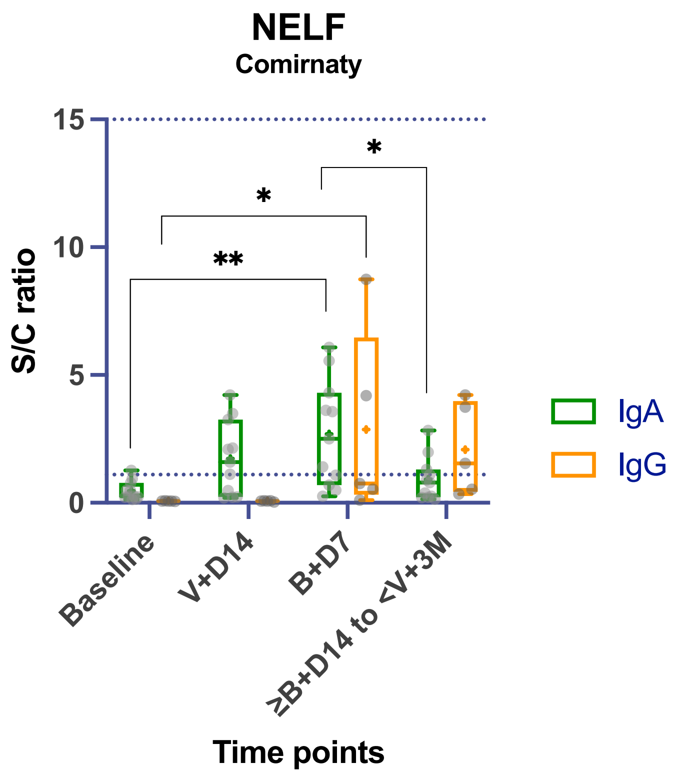


**Fig S3. Dynamic of SARS-CoV-2 S1-specific IgA and IgG in NELFs in extended time point.** 11 subjects received Comirnaty reached the 4$^{th}$ sampling time point. The S/C ratio of the S1-specific IgA was significantly lower in the 4$^{th}$ sampling time point compared to 7±2 days after booster (B+D7), Friedman test ($p$ = 0.018).
